# Microstructural white matter disruptions and their clinical correlates in Wilson disease: A neurite orientation dispersion and density imaging study

**DOI:** 10.64898/2026.03.27.26349503

**Authors:** Ann Carolin Hausmann, Silja K. Querbach, Christian Rubbert, Alfons Schnitzler, Julian Caspers, Christian Johannes Hartmann

**Affiliations:** Institute of Clinical Neuroscience and Medical Psychology, Medical Faculty and University Hospital Düsseldorf, Heinrich-Heine-University Düsseldorf, Moorenstr. 5, 40225 Düsseldorf, Germany; Department of Diagnostic and Interventional Radiology, Medical Faculty and University Hospital Düsseldorf, Heinrich-Heine-University Düsseldorf, Moorenstr. 5, 40225 Düsseldorf, Germany; Department of Neurology, Medical Faculty and University Hospital Düsseldorf, Heinrich-Heine-University Düsseldorf, Moorenstr. 5, 40225 Düsseldorf, Germany

**Keywords:** WD, diffusion MRI, NODDI, DTI, microstructure, free water

## Abstract

**Background:** Neurite orientation dispersion and density imaging (NODDI) shows promise in providing specific insights into the neurite morphology underlying white matter (WM) damage in neurodegenerative diseases. This study aimed to advance the currently limited knowledge by characterizing NODDI-derived microstructural WM alterations in Wilson disease (WD) and examining their relationships with clinical symptoms.

**Methods:** 30 WD patients, including 19 with predominant neurological involvement (neuro-WD) and 11 with hepatic manifestation (hep-WD), and 30 matched healthy controls underwent multi-shell diffusion-weighted magnetic resonance imaging. NODDI metrics, including neurite density index (NDI), orientation dispersion index (ODI), and isotropic volume fraction (ISOVF), and diffusion tensor imaging-based fractional anisotropy (FA) were estimated. Group differences in diffusion parameters across the WM skeleton were determined using tract-based spatial statistics. Additionally, voxel-wise correlations with neurological and cognitive scores were investigated.

**Results:** We observed widespread NDI and ODI reductions in neuro-WD patients and ISOVF increases in hep-WD patients compared with healthy controls, particularly involving the corpus callosum, corona radiata, superior longitudinal fasciculus, external and internal capsule, and superior fronto-occipital fasciculus. A comparable yet more subtle pattern was found when comparing phenotypes. Distinct NDI and ODI constellations were identified as the microstructural determinants of FA alterations. Decreased NDI in the aforementioned fibers were correlated with neurological impairment, processing speed, and visual attention.

**Conclusions:** Phenotype-specific microstructural WM alterations were identified, characterized by globally reduced axonal density and fiber organization in neuro-WD and excess free water in hep-WD. NODDI could be useful as an imaging biomarker for forecasting conversion to neurological WD manifestations and monitoring of disease progression.

## 1. Introduction

Wilson disease (WD) is a rare autosomal-recessive disorder of impaired copper metabolism that leads to pathological copper accumulation in the liver and brain, and may result in hepatic dysfunction, neurological impairment, and cognitive deficits [1]. The most affected cognitive domains are memory, attention, processing speed, verbal fluency, and executive function [2–5]. Neurological manifestations in WD are associated with poorer prognosis and significantly reduced health-related quality of life compared with the hepatic form of the disease, largely attributable to cognitive decline [6,7]. Cerebral copper accumulation induces neurotoxic damage in the brain, primarily affecting deep gray matter nuclei and white matter (WM) tracts [7,8]. While a growing body of research has documented the clinical significance of basal ganglia impairments in WD [9–12], only a limited number of studies have investigated the relationship between WM integrity and clinical symptoms [4,13,14]. Thus, the role of WM alterations in WD-related neurological and cognitive impairment remains poorly understood.

The evaluation of WM in WD is predominantly based on diffusion tensor imaging (DTI) [15], which characterizes brain microstructure using diffusion-weighted magnetic resonance imaging (MRI) by quantifying the directional motion of water molecules [16]. These studies have reported widespread disruption in fiber integrity in WD, reflected by reduced fractional anisotropy (FA) and increased mean diffusivity, measuring the overall directionality and magnitude of water diffusion, respectively [13,14,17,18]. Despite the high sensitivity of DTI metrics to WM damage, their limited biological specificity leaves the underlying pathomechanisms unclear [19]. In addition, a recent study demonstrated increased free water content in the WM of WD patients and a marked attenuation of DTI abnormalities following free water correction [20]. Free water contamination, arising from partial volume effects between, e.g., cerebrospinal fluid and brain parenchyma within a voxel, may bias DTI measurements and contribute to inconsistencies in DTI findings across WD studies, particularly regarding the direction of FA changes [15]. Given its lack of specificity and susceptibility to partial volume effects, especially in regions with complex fiber architecture or inter-tissue interactions, [21,22] conventional DTI may be insufficient to fully characterize WM pathologies in WD.

Recent advancements in diffusion MRI modeling have introduced neurite orientation dispersion and density imaging (NODDI) [23] as a promising candidate for the development of next-generation imaging biomarkers of neuronal microstructure impairment [24,25]. In contrast to the single-tensor model used in DTI, NODDI decomposes the diffusion signal into three distinct tissue compartments [23]. Thereby, specific estimates of neurite density, neurite orientation dispersion, and free water proportion are derived, which are expected to provide biologically more meaningful information [23,24]. Application of NODDI has yielded encouraging results in clinical research, demonstrating high sensitivity and plausible interpretations of disease-related effects, as well as robust correlations with disease severity in patients with Alzheimer’s or Parkinson’s disease [24]. Preliminary NODDI studies in WD patients indicate significant microstructural impairment in the basal ganglia, thalamus, [9,10] and WM [26]. However, the current evidence is scarce, particularly with respect to phenotype-specific changes, and the relationship between NODDI-derived WM metrics and clinical symptoms has not yet been investigated in WD.

With this study, we aimed to advance the currently limited knowledge by characterizing NODDI-derived microstructural WM alterations in WD patients with distinct clinical phenotypes and investigating their relationships with neurological and cognitive symptoms.

## 2. Methods

### 2.1 Study participants

In this prospective, cross-sectional study, we recruited 36 WD patients from the WD outpatient clinic of the Department of Neurology of the University Hospital Düsseldorf, Germany. In addition, 30 age- and sex-matched healthy controls were enrolled. We included patients aged 18 years or older with a Leipzig score of ≥ 4, suggesting an established diagnosis of WD according to international consensus criteria [27]. Patients who initially presented with or developed significant neurological impairment before study enrollment were classified as having a neurological phenotype (neuro-WD). Those without significant neurological impairment at any stage of the disease were classified as having a hepatic phenotype (hep-WD). The exclusion criteria encompassed contraindications to MRI, as well as the presence or history of any neurological disease other than WD or other severe medical conditions that would impede the completion of study assessments. Participants were also examined in a previous study that focused on investigating neuroimaging biomarkers related to gray matter atrophy [28]. In contrast, the current study evaluated microstructural alterations in global WM.

### 2.2 Clinical assessments

Neurological symptoms were thoroughly assessed using the Unified Wilson’s Disease Rating Scale neurological subscale (UWDRS-N) by a senior neurologist in the field of movement disorders (CJH, 15 years of experience with WD). Additionally, standardized neuropsychological testing of multiple cognitive domains was conducted. The Montreal Cognitive Assessment (MoCA) was administered to evaluate global cognitive function. Sustained attention and processing speed were tested using the oral version of the Symbol Digit Modalities Test (SDMT) and the written form of the Trail Making Test Part A (TMTA). Executive function was tested using the Trail Making Test Part B (TMTB). Verbal memory was assessed using the Word List Memory Test list learning (WLMT-L) and delayed recall (WLMT-R) of the Consortium to Establish a Registry for Alzheimer’s Disease (CERAD) test battery and the Semantic Fluency Test (SFT, animals). Cognitive scores were standardized via *z*-transformations using normative data.

### 2.3 Imaging acquisition

High-resolution structural and multi-shell diffusion MRI data were acquired on a 3T MRI scanner (MAGNETOM Prisma, Siemens Healthineers, Erlangen, Germany) using a 64-channel head coil. The protocol included the following sequences and parameters, which were adapted from the Lifespan Human Connectome Project in Aging [29]: 3D T1-weighted magnetization-prepared rapid gradient-echo (MPRAGE) sequence (repetition time [TR] = 2,500 ms, echo time [TE] = 2.22 ms, inversion time [TI] = 1,000 ms, flip angle = 8°, field of view [FoV] = 240*256 mm, slice thickness = 0.8 mm) and diffusion-weighted spin-echo echo-planar imaging (EPI) (TR = 3,230 ms, TE = 89.2 ms, FoV = 210*210 mm, slice thickness = 1.5 mm, multi-band factor 3; 98 directions; *b*-values = 0, 1,500, and 3,000 s/mm^2^). Foam padding was used to minimize head motion. When significant motion artifacts were identified, sequences were repeated within the same scanning session.

### 2.4 Processing of MRI data

The imaging data was preprocessed using the Human Connectome Project minimal preprocessing pipelines (version 4.2.0) [30] employing FreeSurfer (version 5.3) and the Oxford Centre for Functional MRI of the Brain (FMRIB) Software Library (FSL; version 6.0.2) [31].

Structural images were corrected for gradient nonlinearity-induced, readout, and bias field distortions, and were linearly and nonlinearly aligned to the Montreal Neurological Institute (MNI) 152 standard space and the anterior and posterior commissure plane [30]. Undistorted images were skull-stripped to extract brain tissue, followed by segmentation, surface reconstruction, and generation of a structural brain mask [30]. For the diffusion data, b0 intensity was normalized across timeseries and the phase-encoding direction-reversed b0 pairs were used to correct for EPI geometric distortion via FSL’s topup algorithm [32]. Eddy current and head motion correction was performed using the eddy tool [33]. After gradient nonlinearity correction, the mean b0 images were registered to their corresponding T1-weighted images, and diffusion data were resampled and masked with the structural brain masks.

The tensor model was fitted using the dtifit command of FSL’s diffusion toolbox to generate FA maps, which reflect the degree of diffusional directionality, indicative of overall WM integrity. The GPU-accelerated Compute Unified Device Architecture (CUDA) Diffusion Modelling Toolbox (cuDIMOT) [34] within FSL was used to derive the following metrics of the Watson-NODDI model: (1) the neurite density index (NDI), indicating the packing density of axons, (2) the orientation dispersion index (ODI), indicating the spatial organization of axons through variability in fiber orientation, and (3) isotropic volume fraction (ISOVF), indicating free water content [23].

### 2.5 Tract-based spatial statistics

DTI and NODDI maps were analyzed using Tract-Based Spatial Statistics (TBSS) [35]. First, FA maps were registered to the FMRIB58 FA template in the MNI152 standard space using nonlinear transformations. Aligned FA data were averaged across subjects and thinned to generate a study-specific mean WM skeleton, which represents the center of all tracts that are common among our subjects [35]. The skeleton was thresholded at a mean FA of 0.2 to suppress non-WM voxels and those with poor tract correspondence. Each subject’s normalized FA map was projected onto the mean skeleton by filling each point with the maximum value from the center of the nearest relevant tract [35]. The FA-derived nonlinear warps and projections were then applied to skeletonize NDI, ODI, and ISOVF maps in the same manner for subsequent voxel-wise statistical analyses.

We created a one-way analysis of variance (ANOVA) design matrix using the general linear model to compare the diffusion metrics between the neuro-WD, hep-WD, and healthy control groups. An initial *F*-test, followed by pairwise *t*-contrasts was defined. To explore the voxel-wise correlations between diffusion metrics and clinical symptoms in the WD patient cohort, neurological and cognitive scores were added as independent variables. Where significant associations with cognitive scores were identified, subsequent analyses were performed, including UWDRS-N as a covariate. All defined models were controlled for the covariate effects of age and sex. Voxel-wise non-parametric permutation analyses with 5,000 permutations and Threshold-Free Cluster Enhancement (TFCE) were performed using FSL randomise. Results were considered significant at a family-wise error (FWE) corrected *p* ≤ .05. Significant clusters were extracted via the cluster tool in FSL and anatomically localized using the Johns Hopkins University ICBM-DTI-81 WM atlas.

### 2.6 Statistical analysis

Shapiro-Wilk tests were used to evaluate normality. Group differences in demographic data were tested using Fisher-Freeman-Halton exact tests for frequencies and one-way ANOVA for continuous variables. Clinical scores were compared between neuro-WD patients, hep-WD patients, and healthy controls using ANOVAs or Kruskal-Wallis tests, followed by Bonferroni or Dunn’s tests for post hoc pairwise comparisons. Spearman correlation coefficients were used to evaluate the relationship between neurological and neuropsychological scores in the patient cohort. Statistical analyses were performed using SPSS Statistics (v29; IBM Corp, New York) with a level of significance set at *p* ≤ .05. Bonferroni correction was employed to adjust for multiple comparisons.

## 3. Results

### 3.1 Sample demographics and clinical characteristics

Five patients did not complete the MRI acquisition due to technical issues or discomfort, and one patient had an incidental finding. The final sample included 30 WD patients, thereof 19 with neuro-WD and 11 with hep-WD, and 30 healthy controls. Detailed demographic and clinical characteristics are summarized in Table 1. There were no significant differences among the groups in terms of demographic variables, disease duration, or decoppering treatment. None of the participants showed signs of hepatic encephalopathy or had undergone liver transplantation.

**Table 1.** Demographics and clinical characteristics.

|  | Neuro-WD | Hep-WD | Healthy Controls | *p* |
| --- | --- | --- | --- | --- |
| **Demographics** |  |  |  |  |
| *n* | 19 | 11 | 30 | - |
| Male/female ratio | 5/14 | 3/8 | 8/22 | >.999 |
| Age, years | 43.5 (±11.4) | 34.3 (±12.3) | 40.1 (±12.3) | .143 |
| Education, years | 15.6 (±2.5) | 15.2 (±2.7) | 14.8 (±1.9) | .495 |
| Disease duration, years | 24.2 (±11.0) | 17.5 (±9.4) | - | .102 |
| Treatment |  |  | - | .733 |
| D-penicillamine | 8 | 4 |  |  |
| Trientine | 10 | 5 |  |  |
| Chelator and zinc | 1 | 2 |  |  |
| **Neurological score** |  |  |  |  |
| UWDRS-N | 6 [4–15] | 2 [0–4] | 0 [0–0] | **<.001\*\*** |
| **Neuropsychological scores** |  |  |  |  |
| MoCA | 28 [25–29] | 28 [26–28] | 29 [28–29.25] | **.002\*** |
| SDMT, *z* | -0.55 (±0.85) | 0.10 (±0.87) | 0.55 (±0.79) | **<.001\*\*** |
| TMTA, *z* | 0.39 [0.08–0.70] | 0.38 [-0.38–1.15] | 0.42 [0.03–0.84] | .848 |
| TMTB, *z* | 0.59 [0.25–0.78] | 0.24 [-0.35–0.99] | 0.91 [0.50–1.30] | **.031\*** |
| WLMT-L, *z* | -0.28 (±1.38) | -0.42 (±1.17) | 0.40 (±0.78) | **.035\*** |
| WLMT-R, *z* | 0.74 [0.21–1.26] | 0.35 [-0.24–0.94] | 1.03 [0.35–1.41] | .154 |
| SFT, *z* | 0.12 (±1.0) | -0.17 (±1.30) | 0.42 (±0.84) | .222 |

*Notes.* Data is either presented as counts, mean (±standard deviation), or median [interquartile range]. Significant group differences are highlighted in bold, with \* = *p*-values < .05 and \*\* = *p*-values < .001

Patients with neuro-WD demonstrated significantly higher UWDRS-N scores than both hep-WD patients (*p* = .004) and healthy controls (*p* < .001). Based on UWDRS-N subscores [36], the most prevalent neurological symptoms were parkinsonism (68%), tremor (53%), and ataxia (42%), followed by dysarthria (32%), impaired handwriting (32%), dystonia (26%), rigidity (26%), and chorea (5%). MoCA scores were significantly lower in both neuro-WD patients (*p* = .006) and hep-WD patients (*p* = .026) compared with healthy controls. Additionally, patients with neuro-WD performed worse than healthy controls on the SDMT (*p* < .001). Although a significant difference in TMTB and WLMT-L scores was found across the three groups, no specific pairwise differences were identified in post hoc analyses after correcting for multiple testing. No significant differences in TMTA, WLMT-R, and SFT performances were observed between groups. Except for the SDMT (*rs* = -.52, *p* = .023), we found no correlations between neuropsychological and UWDRS-N scores in WD patients.

### 3.2 Differences in diffusion metrics: Neuro-WD patients vs. healthy controls

*F*-Tests revealed significant differences in all diffusion metrics among at least one pair of groups (all FWE-corrected *p* ≤ .05). Subsequent pairwise comparisons yielded significant alterations in NDI, ODI, and FA in neuro-WD patients within extensive, symmetrical WM regions compared with healthy controls (all FWE-corrected *p* ≤ .05; see Fig. 1).

**Fig. 1.**
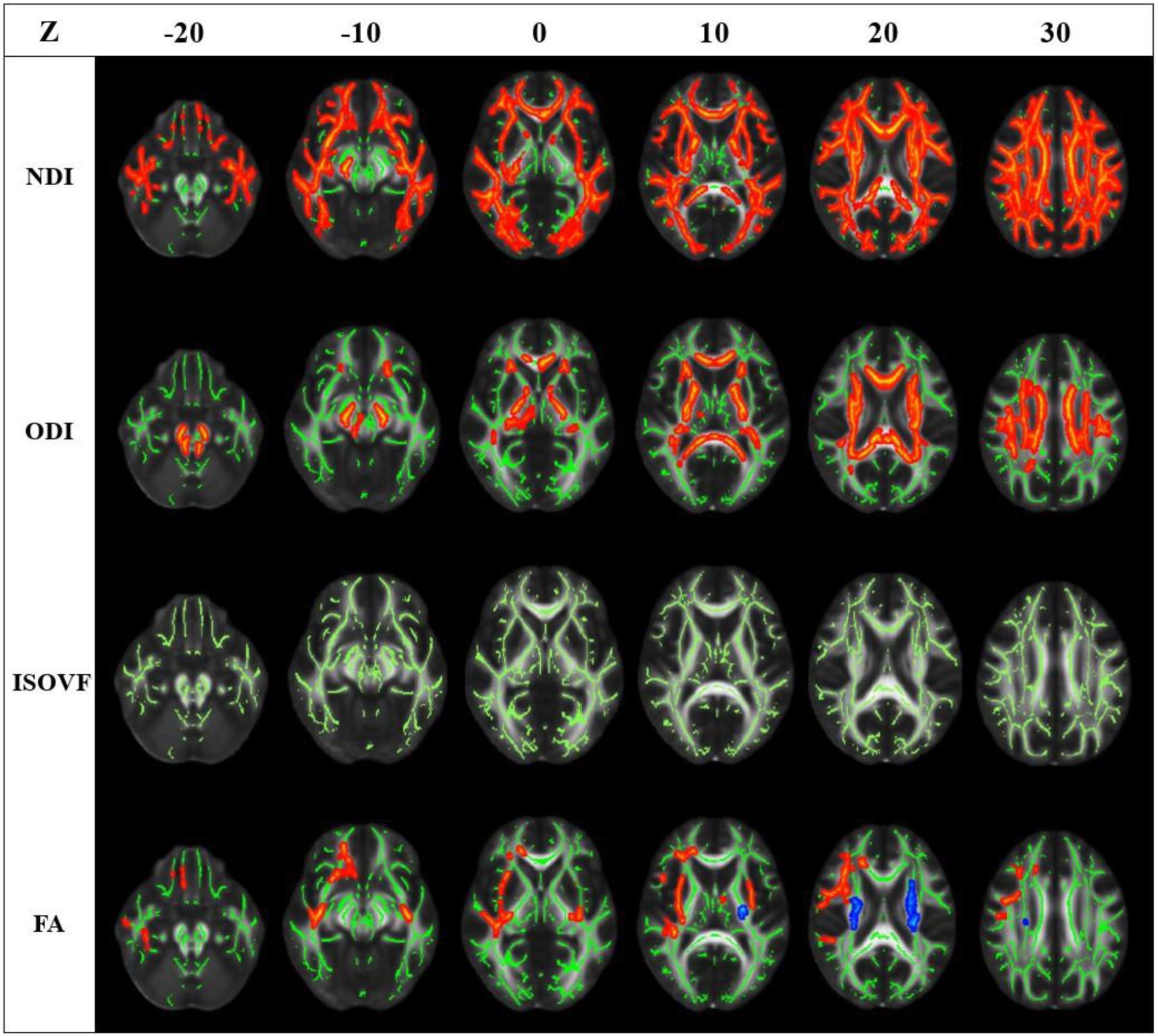
Differences in diffusion metrics between neuro-WD patients and healthy controls. WM regions with significant clusters of decreased (red-yellow) or increased (blue) diffusion metrics in neuro-WD patients compared with healthy controls are shown on the study-specific mean WM skeleton (green) on the FMRIB58 FA template (TFCE, FWE-corrected *p* ≤ .05). Results are presented on representative axial slices, ranging from Z = −20 to 30 (in MNI coordinates). The TBSS fill script was used to aid visualization. Images are displayed in accordance with radiological convention, meaning that the left side of the images corresponds to the right hemisphere of the brain and vice versa

For NDI and ODI, these regions involve the corpus callosum as well as the bilateral superior and anterior corona radiata, superior longitudinal fasciculus, external and internal capsule, posterior thalamic radiation, and superior fronto-occipital fasciculus. Additional NDI reductions were observed within the cingulum and sagittal stratum. In turn, ODI was also reduced within the cerebral peduncles, middle and superior cerebellar peduncles, bilateral corticospinal tracts, and medial lemniscus. In addition, significant FA decreases were found within the bilateral external capsule and right anterior corona radiata, retrolenticular part of internal capsule, superior longitudinal fasciculus, and sagittal stratum. FA increases were restricted to the bilateral superior corona radiata and left posterior limb of internal capsule. While decreases in FA were primarily accompanied by reductions in NDI, rather than changes in ODI, increases in FA were predominantly observed in WM areas that demonstrated parallel reductions in NDI and ODI. Cluster-specific statistics of the comparison between neuro-WD patients and healthy controls are provided in Supplementary File 1 (see Tables S1, S2, and S3).

### 3.3 Differences in diffusion metrics: Hep-WD patients vs. healthy controls

Patients with hep-WD showed no differences in NDI, ODI, or FA metrics compared with healthy controls, but a significant increase in ISOVF within the corpus callosum, bilateral anterior and superior corona radiata, bilateral superior longitudinal fasciculus, left external and internal capsule, and sagittal stratum (all FWE-corrected *p* ≤ .05; see Fig. 2 and Table S4).

**Fig. 2.**
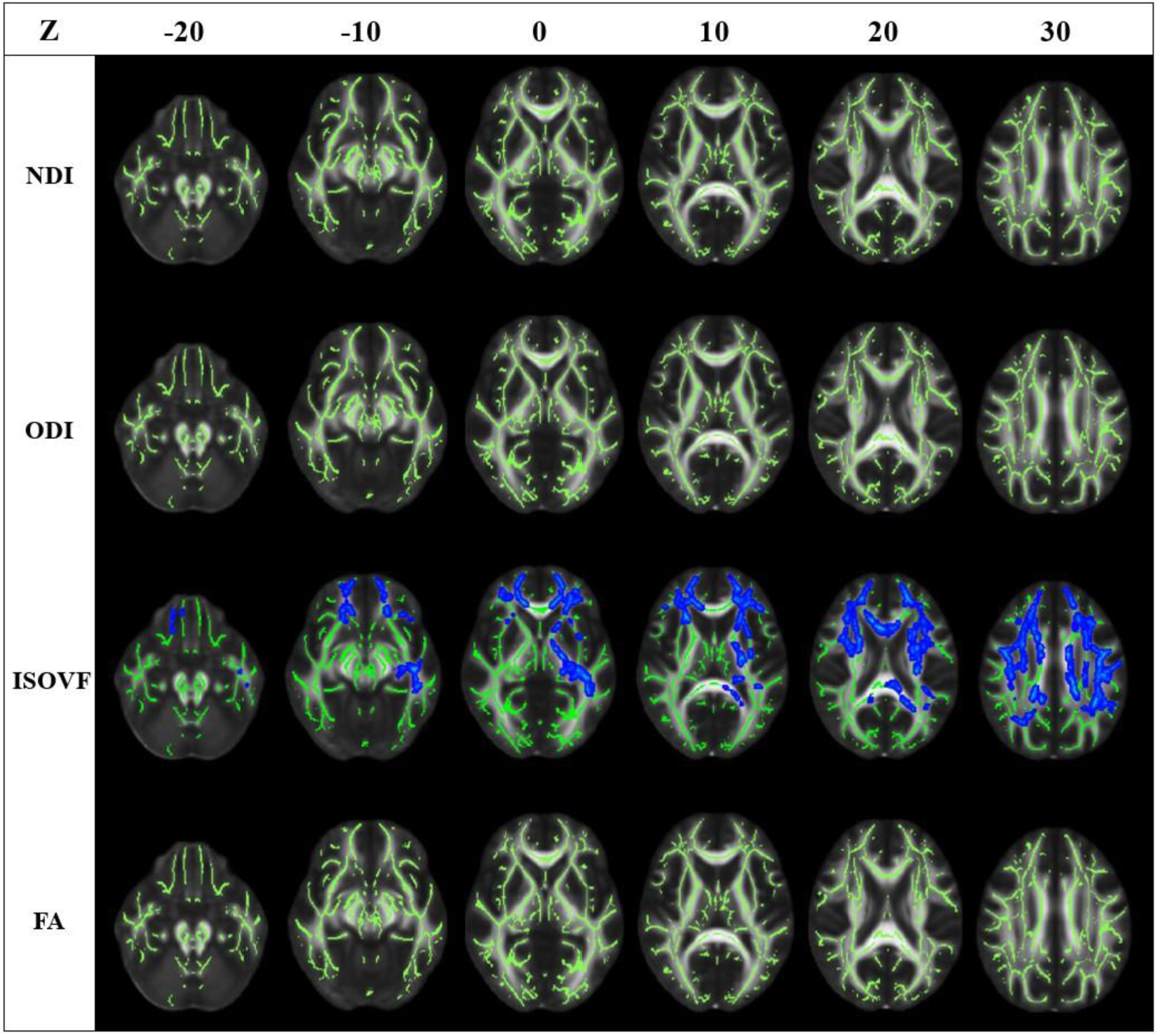
Differences in diffusion metrics between hep-WD patients and healthy controls. WM regions with significant clusters of increased ISOVF (blue) in hep-WD patients compared with healthy controls are shown on the study-specific mean WM skeleton (green) on the FMRIB58 FA template (TFCE, FWE-corrected *p* ≤ .05). Results are presented on representative axial slices, ranging from Z = −20 to 30 (in MNI coordinates). The TBSS fill script was used to aid visualization. The images are displayed in accordance with radiological convention, meaning that the left side of the images corresponds to the right hemisphere of the brain and vice versa

### 3.4 Differences in diffusion metrics: Neuro-WD vs. hep-WD patients

When comparing WD phenotypes, we found that patients with neuro-WD demonstrated significant reductions in NDI and ODI, while those with hep-WD had increased ISOVF (all FWE-corrected *p* ≤ .05; see Fig. 3). The affected WM regions overlapped with those identified in the comparisons with healthy controls, albeit the differences were less pronounced, particularly in NDI. The cluster-specific statistics of the WD subgroup analysis are provided in Supplementary File 1 (see Tables S5, S6, and S7).

**Fig. 3.**
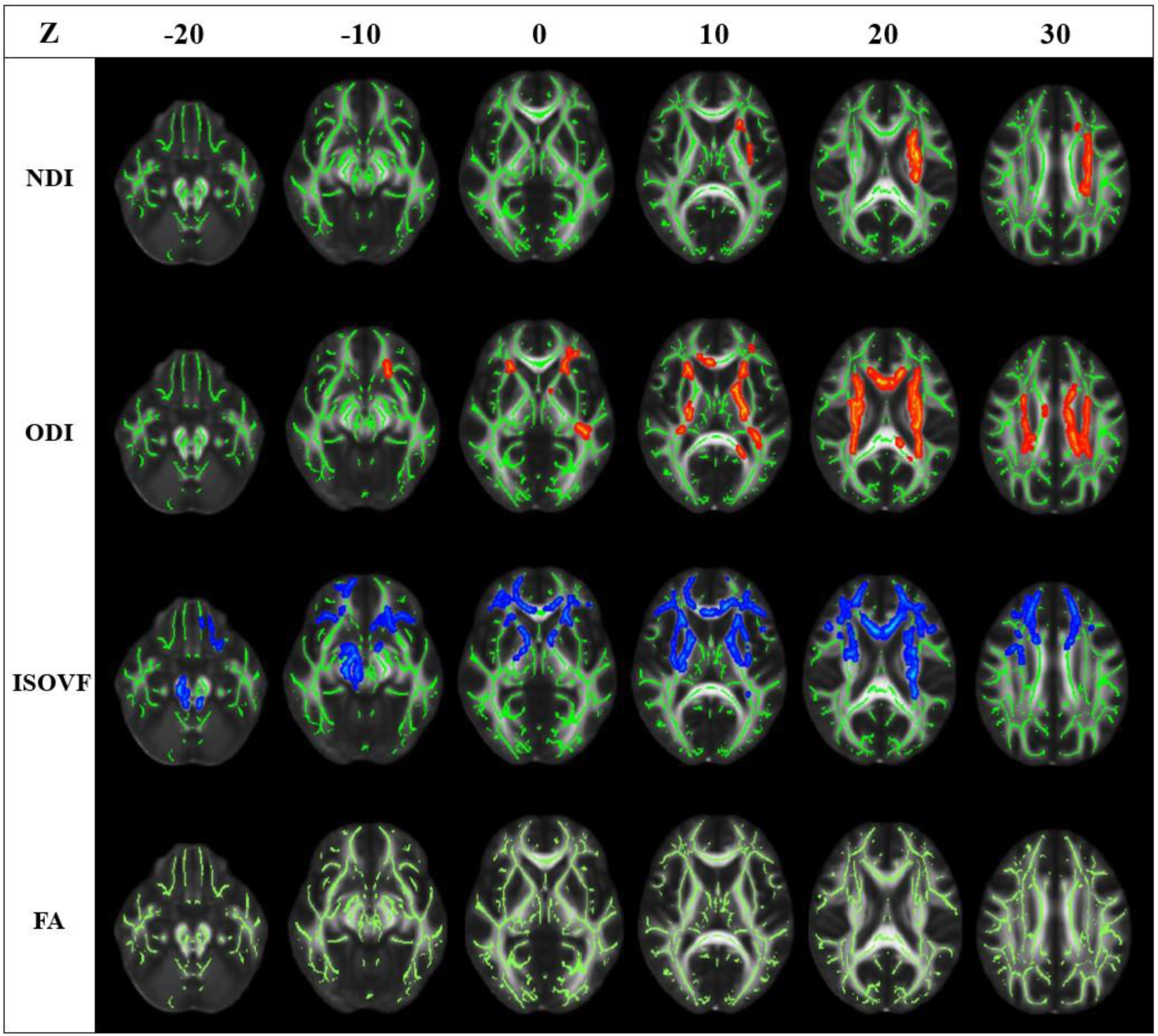
Differences in diffusion metrics between neuro-WD and hep-WD patients. WM regions with significant clusters of decreased diffusion metrics (red-yellow) in neuro-WD patients or increased diffusion metrics (blue) in hep-WD patients are shown on the study-specific mean WM skeleton (green) on the FMRIB58 FA template (TFCE, FWE-corrected *p* ≤ .05). Results are presented on representative axial slices, ranging from Z = −20 to 30 (in MNI coordinates). The TBSS fill script was used to aid visualization. The images are displayed in accordance with radiological convention, meaning that the left side of the images corresponds to the right hemisphere of the brain and vice versa

### 3.5 Correlations with clinical scores

Increased UWDRS-N scores were significantly correlated with NDI reductions within the corpus callosum, bilateral corona radiata, superior longitudinal fasciculus, external and internal capsule, posterior thalamic radiation, corticospinal tract, bilateral superior and middle cerebellar peduncles, and pontine crossing tract (all FWE-corrected *p* ≤ .05; see Fig. 4 and Table S8). Furthermore, poorer performance on the TMTA was significantly correlated with NDI reductions within the corpus callosum, bilateral corona radiata, superior longitudinal fasciculus, external and internal capsule, sagittal stratum, posterior thalamic radiation, fornix, and uncinate fasciculus (controlled for UWDRS-N scores; all FWE-corrected *p* ≤ .05; see Fig. 4 and Table S9). Reduced FA was significantly correlated with increased UWDRS-N scores, as well as poorer TMTA performance in most of the aforementioned WM regions, albeit to a lesser extent (all FWE-corrected *p* ≤ .05; see Fig. 4, Tables S12 and S13). We also identified WM tracts where NDI was decreased when performances on the SDMT or TMTB were diminished (see Fig. S1, Tables S10 and S11), and where FA decreased with poorer performance on the TMTB (see Fig. S2 and Table S14). However, once UWDRS-N scores were included as a covariate, these correlations were no longer significant. No associations between ODI/ISOVF and clinical scores, nor between NODDI metrics and the remaining neuropsychological scores were detected.

**Fig. 4.**
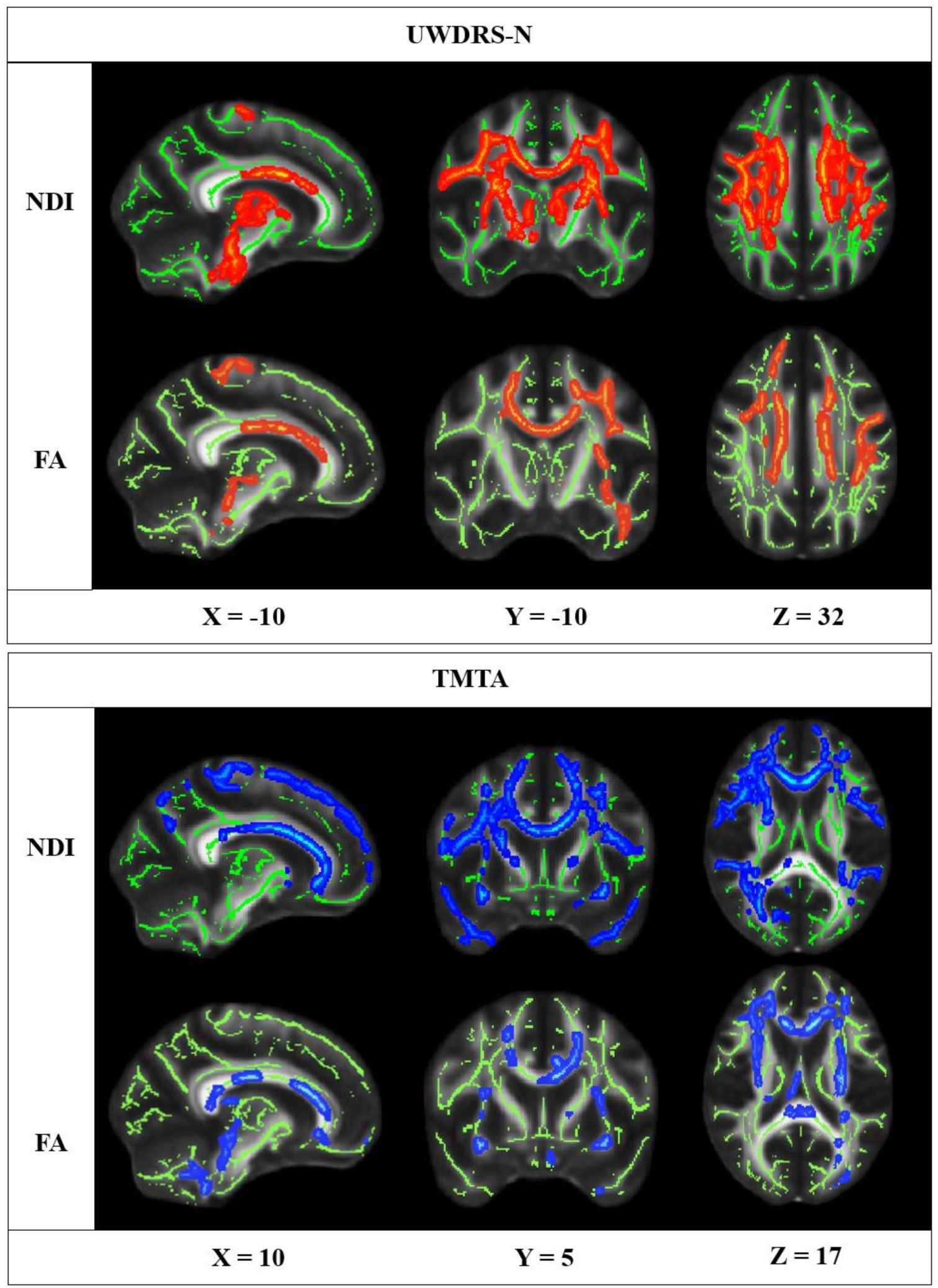
Significant associations between diffusion metrics and clinical scores. WM regions with significant clusters of decreased diffusion metrics with increasing UWDRS-N scores (red-yellow) or decreasing TMTA performance (blue; controlled for UWDRS-N scores) in WD patients are shown on the study-specific mean WM skeleton (green) on the FMRIB58 FA template (TFCE, FWE-corrected *p* ≤ .05). Results are presented on three representative slices (in MNI coordinates). The TBSS fill script was used to aid visualization. The images are displayed in accordance with radiological convention, meaning that the left side of the images corresponds to the right hemisphere of the brain and vice versa

## 4. Discussion

This study systematically investigated WM alterations and their association with neurological and cognitive symptoms in patients with WD. We provide initial evidence for phenotype-specific differences in NODDI-derived WM microstructure, characterized by increased ISOVF in hep-WD patients and widespread reductions in NDI and ODI in neuro-WD patients. Notably, NDI decreases across several major WM tracts were further identified as neuroimaging correlates of neurological impairment, processing speed, and visual attention. Therefore, this study offers novel insights into pathophysiological processes and underlines the clinical relevance of WM impairment in WD.

The observed NDI decreases in patients with neuro-WD resonate well with the preliminary results of Song et al., who reported widely reduced NDI in WD patients within WM regions that correspond to those identified in our study, including the corpus callosum, bilateral corona radiata, superior longitudinal fasciculus, superior fronto-occipital fasciculus, and thalamic radiation [26]. In contrast to their conference report, we additionally identified significant ODI reductions in these pathways and the brainstem. Microstructural abnormalities in the aforementioned fibers have also been identified in other neurodegenerative disorders with symptoms overlapping those of WD, such as tremor in Parkinson’s disease and chorea in Huntington’s disease [19]. Our results suggest global reductions in axonal density and a simplified fiber organization in neuro-WD patients compared with healthy controls. These observations are consistent with postmortem evidence of axonal loss, demyelination, cavitation, and tissue spongiosis in WD [7,37], thereby supporting the biological plausibility of our findings.

Furthermore, we identified significantly increased ISOVF across diffuse WM regions in patients with hep-WD, which were previously only reported in the corpus callosum of WD patients [26]. Given that NDI and ODI did not differ between hep-WD patients and healthy controls, indicating the preservation of broader tissue integrity, this excess free water is unlikely to be attributable to atrophy. A similar perspective is shared by Jing et al., who recently observed an increase in free water using bi-tensor free water imaging in hep-WD patients in WM regions overlapping with those identified in our study, including the corpus callosum, superior longitudinal fasciculus, anterior limb of internal capsule, and large parts of the corona radiata [20]. The DTI abnormalities in their hep-WD patients disappeared following free water correction [20], suggesting a closer link to neuroinflammation rather than atrophy. In accordance with this hypothesis, astrocytes initially buffer elevated copper concentrations in the brain, resulting in a disruption in ionic homeostasis, cellular swelling, increased oxidative stress, and the activation of inflammatory responses [1,38]. One of the earliest consequences of cerebral copper overload is, thus, hydropic swelling of astrocytes and myelin sheaths, as well as astrogliosis [1,7]. In light of these considerations, the increased ISOVF observed in this study presumably reflects the presence of subclinical edema or neuroinflammation in the WM of hep-WD patients. The absence of ISOVF alterations in neuro-WD patients may be attributable to the recent observations in molecular studies indicating that chronic copper accumulation exceeds the buffering capacity of astrocytes, resulting in the exhaustion of their neuroprotective function and eventual death through cuproptosis [39]. Nevertheless, another contributing factor to our findings could be persistent hepatic dysfunction and subsequent portosystemic shunting of ammonia, causing neuropathological abnormalities that resemble those induced by copper [1,37]. This viewpoint is challenged by the absence of signs of hepatic encephalopathy in the examination of our patients and a previous NODDI study that did not identify ISOVF changes in patients with minimal hepatic encephalopathy compared with healthy controls [40]. However, it should be noted that the clinical criteria for hepatic encephalopathy are rather broad, meaning that more subtle signs may be missed.

The comparison of WD subgroups yielded a similar, but more subtle, pattern of findings than when each WD group was compared with healthy controls. The extent of significant NDI reductions in neuro-WD patients was far less widespread when compared with hep-WD patients than when compared with healthy controls. This diminished discrepancy was primarily observed in WM regions that showed increased ISOVF in hep-WD patients, suggesting that excess free water in hepatic patients could contribute to progressive axonal injury over time. This hypothesis is consistent with the premise that copper exposure triggers a cascade of inflammatory pathways that ultimately leads to neurodegeneration [41]. Furthermore, the differences between neuro-WD and hep-WD patients in NDI were less pronounced than in ODI, which indicates that axonal loss may be initiated at an earlier stage of cerebral copper deposition than a reduction in axonal complexity. Taken together, our results suggest that NODDI could be a useful tool to assess the transition of WM alterations in WD from a potentially reversible edematous or neuroinflammatory state to axonal degeneration as the disease progresses from a hepatic to a neurological phenotype, even before becoming clinically evident. To clarify the role and temporal trajectory of these changes, longitudinal studies using NODDI should be conducted in patients with different disease states, including treatment-naïve patients, in addition to the assessment of laboratory copper indices.

Consistent with previously reported inconsistencies in FA findings across WD studies [15], we observed FA alterations in both directions in neuro-WD patients. A decrease in FA was primarily observed in WM tracts with decreased NDI but unchanged ODI, while an increase in FA was observed in regions with parallel decreases in NDI and ODI. On the one hand, this finding indicates that the observed reductions in FA were primarily attributable to decreased axonal density and reflect a genuine loss of WM integrity within the bilateral external capsule, as well as the right anterior corona radiata, retrolenticular part of internal capsule, superior longitudinal fasciculus, and sagittal stratum. This aligns with previous studies [14,18,42]. On the other hand, the concomitant presence of reduced axonal density and fiber dispersion may paradoxically increase FA, which can be misinterpreted if ODI is not considered. A similar observation was reported in the WM of Alzheimer’s disease patients by Douaud et al. (2011), who proposed that the misleading FA increase could be attributable to the selective degeneration of individual fiber populations within crossing fibers. This may result in a less diverse pattern in orientation dispersion and, consequently, in an apparently increased alignment of the preserved axons [24,43]. Our study found that FA was increased significantly within the superior corona radiata and the posterior limb of the internal capsule, which are known for their complex fiber architecture [44]. Thus, we recommend that future WD studies examine potential selective damage to crossing fibers within these key regions more thoroughly by employing targeted techniques, such as fixel-based analysis or probabilistic tractography [45,46]. In summary, our findings provide plausible explanations for the bidirectional FA alterations in WD by demonstrating how NODDI can disentangle the respective contributions of axonal loss and changes in fiber organization.

Significant correlations were identified between increased UWDRS-N scores and decreased NDI in motor-related WM tracts. Our findings align with previously reported correlations between neurological symptoms and microstructural alterations within the corticospinal tract [13], left internal capsule, thalamic radiation, corpus callosum [36], and superior cerebellar peduncles in WD [20]. The corpus callosum, the principal WM fiber bundle in the brain, was found to be consistently affected in all our analyses. Callosal microstructural integrity is associated with interhemispheric communication, motor control, and the coordination of bimanual motor tasks [47,48]. In accordance with our findings, a previous study has demonstrated that WD patients with corpus callosum abnormalities exhibited more severe neurological dysfunction compared to patients without such lesions [49]. In addition, we identified a correlation between neurological symptoms and microstructural alterations within the corona radiata and internal capsule, including thalamic radiation and corticospinal tract fibers, which form large, interconnected pathways that link the cerebral cortex, thalamus, and brainstem. Fibers within the internal capsule also traverse the basal ganglia, and fiber volumes between the putamen and thalamus have been demonstrated to be lower in WD patients who present with hypokinesia [50]. Given that the brain regions interconnected by the compromised fibers identified in our study have previously been implicated in the neurological severity of WD [36,51,52], we hypothesize that a reduction in axonal density may further exacerbate motor dysfunction in patients with WD by hampering connectivity between these structures. This would complement the findings of previous studies, which have indicated that extrapyramidal symptoms in WD can be attributable to structural and functional abnormalities within the cerebello-thalamo-cortical network [50,53,54]. Overall, our findings suggest that axonal degeneration in several large WM pathways, such as the corpus callosum, internal capsule, and corona radiata, may contribute to the manifestation of more severe neurological symptoms in patients with WD.

Both WD groups performed worse on the MoCA than healthy controls, and neuro-WD patients also had significantly lower SDMT scores. We additionally found group differences in TMTB and WLMT-L performances that could not be further resolved in post hoc analyses, presumably due to the limited statistical power resulting from small sample sizes. Our findings align with those of earlier studies that reported a mild decline in global cognitive function, attention, processing speed, executive function, and verbal learning in patients with WD [2–5]. Overall, the microstructural alterations identified in this study also compromised WM fibers integral to high-level cognitive functions. In addition to its role in motor function, the corpus callosum is considered essential to executive and global cognitive function, with the genu contributing significantly to working memory [55]. Likewise, the microstructural integrity of the corona radiata has been linked to multiple dimensions of attention in healthy individuals [56], while the superior longitudinal fasciculus plays a critical role in integrating information underlying complex cognitive functions, including language processing, visuospatial attention, verbal working memory, and global executive function [55,57]. In line with these considerations, we identified significant correlations between reduced NDI in these regions and poorer performances on the TMTA, TMTB, and SDMT. However, only the correlation with TMTA performance persisted after controlling for UWDRS-N scores, suggesting that subtle dysarthria and mildly impaired handwriting ability may have confounded TMTB and SDMT performance through a reduction in motor speed. Therefore, our findings indicate that regional reductions in axonal density affect processing speed, visual attention, and/or visuomotor coordination in patients with WD. This aligns with the observation that, while multifocal cerebral pathologies tend to negatively affect cognitive performance in WD, they do not necessarily result in a significant deterioration of cognitive deficits beyond the impact of basal ganglia impairments [5]. Moreover, given that no correlations were found between clinical scores and ODI or ISOVF, the observed changes in fiber dispersion and free water probably did not impact neurological or cognitive function in our patients.

The present findings may have several implications. While both DTI and NODDI demonstrated WM impairment in neuro-WD patients, the extent of NODDI alterations was found to be more pronounced and plausibly explained the specific microstructural underpinnings of bidirectional FA scalars. In addition, the identified increase in free water fraction in hep-WD patients can bias DTI-derived conclusions due to the model’s susceptibility to partial volume contamination. Therefore, we recommend the implementation of advanced diffusion MRI modeling with NODDI to complement DTI in future studies to more accurately assess WM microstructure in WD. In the past, DTI metrics have been proposed as useful markers in earlier stages of WD [15], as they can trace brain abnormalities prior to the manifestation of morphological changes in structural MRI and clinically overt neurological symptoms [42] and may improve following decoppering therapy [58]. This study presents compelling evidence that NODDI could offer additional value in the clinical evaluation of WD through its apparent sensitivity to different progression stages of copper overload, as indicated by its capacity to discern subclinical signs of cerebral copper toxicity in hep-WD as well as long-term axonal disruption in neuro-WD. NDI was identified as a potentially valuable candidate for the development of novel imaging biomarkers, as we demonstrated that decreased axonal density may contribute to WD-related neurological impairment, as well as to lower processing speed and visual attention. Future longitudinal studies may offer further insights into the benefits of using NODDI-derived biomarkers in MRI evaluations to potentially assist in disease monitoring to adjust treatment regimens before clinically meaningful axonal degeneration manifests.

This study has several limitations. The limited sample size constrained the statistical power; however, given the very low prevalence of WD, recruitment of eligible patients is inherently challenging, and the sample size of the present study is substantial compared with that of other neuroimaging studies in WD. In addition, neuro-WD patients in our study exhibited relatively low UWDRS-N scores, as severe neurological impairments precluded completion of the study assessments. Consequently, microstructural alterations in more severe cases may have been underestimated. Since we focused on leveraging NODDI’s tissue specificity to aid interpretation of FA findings, future studies may benefit from including additional DTI parameters, such as mean diffusivity. We further decided to employ the standard Watson-NODDI model, which has demonstrated reliable performance in clinical research [24] and in initial WD studies [9,10]. Nevertheless, subsequent studies could extend these analyses by implementing the Bingham-NODDI extension, which additionally quantifies the anisotropic orientation dispersion of neurites [59]. Finally, although there is increasing evidence for the potential of advanced diffusion MRI modeling, translating these novel techniques into clinical practice remains challenging and will require further validation.

In conclusion, this study has revealed phenotype-specific microstructural WM alterations in patients with WD, characterized by excess free water in hep-WD and global reductions in axonal density and fiber organization in neuro-WD. Our results suggest that NODDI could be a viable method for forecasting the conversion from edematous or neuroinflammatory states in hep-WD patients to clinically meaningful WM neurodegeneration in neuro-WD patients. Decreased neurite density primarily contributed to the severity of neurological impairment but also negatively affected processing speed and visual attention. Overall, NODDI-derived diffusion metrics may prove useful for monitoring disease progression and could serve as potential imaging biomarkers for the early detection of the conversion to neurological manifestations in WD.

## 5. Declarations

### 5.3 Ethical Statements

This prospective, cross-sectional study was part of a larger observational imaging study on neurodegenerative diseases (DRKS00034050). All procedures were performed in compliance with relevant laws, institutional guidelines, and the principles put forth in the Declaration of Helsinki in its latest revision (October 2024) and have been approved by the local ethics committee of the Medical Faculty at the Heinrich-Heine-University Düsseldorf, Germany (2019-470_4; 7th of February 2022). Written informed consent was obtained from all subjects prior to study participation. The privacy rights of all participants have been observed.

### 5.4 Data availability statement

Study data is not publicly available due to privacy reasons, but anonymized data may be requested from the corresponding author upon reasonable request.

### 5.5 Authors’ contribution statements

Ann Carolin Hausman was involved in the study conception, data collection, and data curation, performed all formal analyses, visualized the results, wrote and revised the first draft, and prepared the final manuscript. Silja K. Querbach contributed to the study conception and data collection, assisted in methodologies, and commented on previous versions of the manuscript. Christian Rubbert contributed to data quality assurance, provided resources, and critically reviewed the manuscript. Alfons Schnitzler was involved in supervision, provided resources, and critically reviewed the manuscript. Julian Caspers was involved in the study conception, data quality assurance, provided resources, and critically reviewed the manuscript. Christian Johannes Hartmann contributed to the study conception, data collection, supervision, and critically reviewed the first draft and the final manuscript. All authors read and approved the final manuscript.

## Supplementary material

Hausmann et al_Supplementary File 1: This supplementary material contains the cluster-specific statistics of the significant tract-based spatial statistics results and additional figures of the correlation analyses.

## Acknowledgements

We would like to sincerely thank all the participants who dedicated their time to contributing to this study.

