## Supplementary File 1 for "Microstructural white matter disruptions and their clinical correlates in Wilson disease: A neurite orientation dispersion and density imaging study"

### 1. Cluster-specific statistics: Group differences in diffusion metrics

In all supplementary tables, the peak MNI coordinates, total voxels, and minimum *p*-value of significant clusters are shown, followed by the JHU atlas-derived localization of included WM regions with their voxel count and mean *p*-values (FWE-corrected). To mitigate the occurrence of false positives, only robust clusters and regions with an overlap of more than 100 voxels are displayed.

WM = white matter; NDI = neurite density index; ODI = orientation dispersion index; ISOVF = isotropic volume fraction; FA = fractional anisotropy; neuro-WD = Wilson disease patients with a neurological phenotype; hep-WD = Wilson disease patients with a hepatic phenotype; MNI = Montreal Neurological Institute; JHU = Johns Hopkins University; B = body; G = genu; S = splenium; R = right; L = left; M = middle; FWE = family-wise error; UWDRS-N = Unified Wilson's Disease Rating Scale neurological subscale; TMTA = Trail Making Test Part A; TMTB = Trail Making Test Part B; SDMT = Symbol Digit Modalities Test; TBSS = Tract-Based Spatial Statistics; TFCE = Threshold-Free Cluster Enhancement.

#### 1.1 Neuro-WD patients vs. healthy controls

**Table S1 WM regions with decreased NDI in neuro-WD patients compared with healthy controls**

| Coordinates | | | WM regions | Side | Voxels | Total voxels | Mean *p* |
|---|---|---|---|---|---|---|---|
| X | Y | Z | | | | | |
| 35 | 5 | -2 | | | | 77,129 | $p_{min} < .001$ |
| | | | Corpus callosum | B | 2,758 | 5,391 | .008 |
| | | | | G | 1,465 | | |
| | | | | S | 1,168 | | |
| | | | Superior corona radiata | R | 1,455 | 2,829 | .002 |
| | | | | L | 1,374 | | |
| | | | Superior longitudinal fasciculus | R | 1,469 | 2,762 | .005 |
| | | | | L | 1,293 | | |
| | | | Anterior corona radiata | R | 1,298 | 2,613 | .006 |
| | | | | L | 1,315 | | |
| | | | External capsule | R | 1,014 | 2,076 | .006 |

| | | | | |
|---|---|---|---|---|
| | L | 1,062 | | |
| Posterior corona radiata | R | 587 | 1,129 | .008 |
| | L | 542 | | |
| Posterior thalamic radiation | R | 604 | 959 | .020 |
| | L | 355 | | |
| Anterior limb of internal capsule | R | 448 | 871 | .006 |
| | L | 423 | | |
| Retrolenticular part of internal capsule | R | 429 | 722 | .012 |
| | L | 293 | | |
| Posterior limb of internal capsule | R | 446 | 710 | .010 |
| | L | 264 | | |
| Cingulum | R | 260 | 645 | .013 |
| | L | 385 | | |
| Sagittal stratum | R | 319 | 593 | .014 |
| | L | 274 | | |
| Superior fronto-occipital fasciculus | R | 106 | 203 | .002 |
| | L | 97 | | |
| Fornix | R | 163 | 196 | .012 |
| | L | 33 | | |
| Cerebral peduncle | R | 175 | | .011 |
| Uncinate fasciculus | R | 75 | 132 | .007 |
| | L | 59 | | |

**Table S2 WM regions with decreased ODI in neuro-WD patients compared with healthy controls**

| Coordinates | | | WM regions | Side | Voxels | Total voxels | Mean *p* |
|---|---|---|---|---|---|---|---|
| X | Y | Z | | | | | |
| -12 | -18 | -16 | | | | 20,736 | $p_{min} < .001$ |
| | | | Corpus callosum | B | 2,552 | 5,852 | .003 |
| | | | | G | 1,275 | | |
| | | | | S | 2,025 | | |
| | | | Superior corona radiata | R | 1,062 | 1,991 | .002 |
| | | | | L | 929 | | |
| | | | Posterior limb of internal capsule | R | 640 | 1,362 | .002 |
| | | | | L | 722 | | |
| | | | Superior longitudinal fasciculus | R | 634 | 932 | .006 |
| | | | | L | 298 | | |
| | | | Anterior corona radiata | R | 481 | 922 | .003 |
| | | | | L | 441 | | |
| | | | | R | 411 | 834 | .003 |

| | WM regions | Side | Voxels | Total voxels | Mean p |
|---|---|---|---|---|---|
| | Anterior limb of internal capsule | L | 423 | | |
| | Posterior corona radiata | R | 424 | 797 | .004 |
| | | L | 373 | | |
| | Cerebral peduncle | R | 404 | 770 | .004 |
| | | L | 366 | | |
| | Cerebellar peduncle | M | 510 | 510 | .020 |
| | Retrolenticular part of internal capsule | R | 247 | 477 | .004 |
| | | L | 230 | | |
| | Corticospinal tract | R | 159 | 394 | .005 |
| | | L | 235 | | |
| | Superior cerebellar peduncle | R | 166 | 322 | .010 |
| | | L | 156 | | |
| | Posterior thalamic radiation | R | 231 | 316 | .007 |
| | | L | 85 | | |
| | External capsule | R | 76 | 225 | .005 |
| | | L | 149 | | |
| | Medial lemniscus | R | 111 | 199 | .013 |
| | | L | 88 | | |
| | Superior fronto-occipital fasciculus | R | 89 | 168 | .002 |
| | | L | 79 | | |

**Table S3 WM regions with altered FA in neuro-WD patients compared with healthy controls**

| **FA decrease** | | | | | | | |
|---|---|---|---|---|---|---|---|
| **Coordinates** | | | **WM regions** | **Side** | **Voxels** | **Total voxels** | **Mean *p*** |
| **X** | **Y** | **Z** | | | | | |
| 32 | 6 | 4 | | | | 4,329 | $p_{min}$ = .016 |
| | | | External capsule | R | 750 | 750 | .027 |
| | | | Anterior corona radiata | R | 511 | 511 | .037 |
| | | | Retrolenticular part of internal capsule | R | 258 | 258 | .033 |
| | | | Superior longitudinal fasciculus | R | 130 | 130 | .036 |
| | | | Sagittal stratum | R | 126 | 126 | .035 |
| -33 | -8 | 6 | | | | 528 | $p_{min}$ = .020 |
| | | | External capsule | L | 413 | 413 | .033 |

| Coordinates | | | WM regions | Side | Voxels | Total voxels | Mean *p* |
|---|---|---|---|---|---|---|---|
| **FA increase** | | | | | | | |
| **X** | **Y** | **Z** | | | | | |
| -24 | -21 | 22 | | | | 532 | $p_{min}$ = .022 |
| | | | Superior corona radiata | L | 258 | 258 | .032 |
| | | | Posterior limb of internal capsule | L | 174 | 174 | .036 |
| 22 | -8 | 18 | | | | 245 | $p_{min}$ = .030 |
| | | | Superior corona radiata | R | 160 | 160 | .036 |

#### 1.2 Hep-WD patients vs. healthy controls

**Table S4 WM regions with increased ISOVF in hep-WD patients compared with healthy controls**

| Coordinates | | | WM regions | Side | Voxels | Total voxels | Mean *p* |
|---|---|---|---|---|---|---|---|
| **X** | **Y** | **Z** | | | | | |
| -19 | 47 | 1 | | | | 10,793 | $p_{min}$ = .011 |
| | | | Corpus callosum | B | 325 | 769 | .030 |
| | | | | G | 55 | | |
| | | | | S | 389 | | |
| | | | Anterior corona radiata | L | 611 | 611 | .021 |
| | | | Superior longitudinal fasciculus | L | 589 | 589 | .022 |
| | | | Superior corona radiata | L | 421 | 421 | .024 |
| | | | Posterior limb of internal capsule | L | 300 | 300 | .041 |
| | | | External capsule | L | 254 | 254 | .030 |
| | | | Anterior limb of internal capsule | L | 205 | 205 | .026 |
| | | | Retrolenticular part of internal capsule | L | 196 | 196 | .042 |
| | | | Fornix | L | 128 | 128 | .042 |
| | | | Posterior corona radiata | L | 122 | 122 | .024 |
| | | | Sagittal stratum | L | 107 | 107 | .041 |

| | | | | | | | |
|---|---|---|---|---|---|---|---|
| 21 | 49 | -7 | | | | 7,283 | $p_{min}$ = .028 |
| | | | Corpus callosum | B | 534 | 950 | .038 |
| | | | | G | 284 | | |
| | | | | S | 132 | | |
| | | | Anterior corona radiata | R | 642 | 642 | .038 |
| | | | Superior corona radiata | R | 519 | 519 | .040 |
| | | | Superior longitudinal fasciculus | R | 397 | 397 | .039 |

##### 1.3 WD subgroups: Neuro-WD patients vs. hep-WD patients

**Table S5 WM regions with decreased NDI in neuro-WD patients compared with hep-WD patients**

| Coordinates | | | WM regions | Side | Voxels | Total voxels | Mean *p* |
|---|---|---|---|---|---|---|---|
| X | Y | Z | | | | | |
| -31 | -11 | 12 | | | | 2,632 | $p_{min}$ = .037 |
| | | | Superior corona radiata | L | 872 | 872 | .040 |
| | | | External capsule | L | 186 | 186 | .041 |

**Table S6 WM regions with decreased ODI in neuro-WD patients compared with hep-WD patients**

| Coordinates | | | WM regions | Side | Voxels | Total voxels | Mean *p* |
|---|---|---|---|---|---|---|---|
| X | Y | Z | | | | | |
| -25 | -10 | 17 | | | | 5,639 | $p_{min}$ < .001 |
| | | | Corpus callosum | B | 1,315 | 2,076 | .019 |
| | | | | G | 474 | | |
| | | | | S | 287 | | |
| | | | Superior corona radiata | L | 802 | 802 | .006 |
| | | | Anterior corona radiata | R | 1 | 502 | .010 |
| | | | | L | 501 | | |
| | | | Posterior limb of internal capsule | L | 434 | 434 | .009 |
| | | | Posterior corona radiata | L | 323 | 323 | .013 |
| | | | Anterior limb of internal capsule | L | 316 | 316 | .012 |

| | | | | | | | |
|---|---|---|---|---|---|---|---|
| | | | Retrolenticular part of internal capsule | L | 292 | 292 | .018 |
| 25 | -10 | 16 | | | | 2,079 | $p_{min}$ = .001 |
| | | | Superior corona radiata | R | 563 | 563 | .011 |
| | | | Anterior corona radiata | R | 457 | 457 | .012 |
| | | | Posterior corona radiata | R | 292 | 292 | .018 |
| | | | Posterior limb of internal capsule | R | 244 | 244 | .011 |
| | | | Retrolenticular part of internal capsule | R | 163 | 163 | .016 |
| | | | Anterior limb of internal capsule | R | 103 | 103 | .017 |

**Table S7 WM regions with increased ISOVF in hep-WD patients compared with neuro-WD patients**

| **Coordinates** | | | **WM regions** | **Side** | **Voxels** | **Total voxels** | **Mean *p*** |
|---|---|---|---|---|---|---|---|
| **X** | **Y** | **Z** | | | | | |
| -13 | 25 | 48 | | | | 13,856 | $p_{min}$ = .016 |
| | | | Corpus callosum | B | 726 | 1,449 | .030 |
| | | | | G | 723 | | |
| | | | Anterior corona radiata | R | 421 | 1,042 | .041 |
| | | | | L | 621 | | |
| | | | Anterior limb of internal capsule | R | 321 | 667 | .037 |
| | | | | L | 346 | | |
| | | | External capsule | R | 221 | 534 | .040 |
| | | | | L | 313 | | |
| | | | Superior corona radiata | R | 284 | 521 | .041 |
| | | | | L | 237 | | |
| | | | Posterior limb of internal capsule | R | 270 | 482 | .040 |
| | | | | L | 212 | | |
| | | | Cerebellar peduncle | M | 337 | 337 | .038 |
| | | | Cerebral peduncle | R | 329 | 329 | .033 |
| | | | Posterior corona radiata | L | 125 | 125 | .039 |

### 2. Cluster-specific statistics: Correlation analyses

#### 2.1 Correlations between NDI and clinical scores

##### Table S8 WM regions where NDI decreased with higher UWDRS-N scores

| Coordinates | | | WM regions | Side | Voxels | Total voxels | Mean *p* |
|---|---|---|---|---|---|---|---|
| X | Y | Z | | | | | |
| 38 | -8 | 32 | | | | 19,128 | $p_{min}$ = .035 |
| | | | Corpus callosum | B | 1,478 | 2,257 | .041 |
| | | | | G | 363 | | |
| | | | | S | 416 | | |
| | | | Superior corona radiata | R | 863 | 1,893 | .040 |
| | | | | L | 1,030 | | |
| | | | Superior longitudinal fasciculus | R | 554 | 1,085 | .042 |
| | | | | L | 531 | | |
| | | | Anterior corona radiata | R | 275 | 791 | .041 |
| | | | | L | 516 | | |
| | | | External capsule | R | 516 | 752 | .041 |
| | | | | L | 236 | | |
| | | | Posterior corona radiata | R | 475 | 727 | .040 |
| | | | | L | 252 | | |
| | | | Posterior limb of internal capsule | R | 338 | 616 | .041 |
| | | | | L | 278 | | |
| | | | Posterior thalamic radiation | R | 176 | 566 | .042 |
| | | | | L | 390 | | |
| | | | Anterior limb of internal capsule | R | 321 | 514 | .041 |
| | | | | L | 193 | | |
| | | | Retrolenticular part of internal capsule | R | 256 | 363 | .042 |
| | | | | L | 107 | | |
| | | | Corticospinal tract | R | 107 | 343 | .044 |
| | | | | L | 236 | | |
| | | | Medial lemniscus | R | 137 | 301 | .040 |
| | | | | L | 164 | | |
| | | | Cerebral peduncle | R | 171 | 253 | .044 |
| | | | | L | 82 | | |
| | | | Pontine crossing tract | | 206 | 206 | .044 |
| | | | Superior cerebellar peduncle | R | 91 | 180 | .041 |
| | | | | L | 89 | | |
| | | | Cerebellar peduncle | M | 125 | 125 | .045 |
| | | | Superior fronto-occipital fasciculus | R | 48 | 118 | .041 |
| | | | | L | 70 | | |
| -30 | -56 | -1 | | | | 427 | $p_{min}$ = .049 |

| | | | | | | | |
|---|---|---|---|---|---|---|---|
| | | | Posterior thalamic radiation | L | 113 | 113 | .049 |

**Table S9 WM regions where NDI decreased with poorer TMTA performance (controlled for UWDRS-N scores)**

| Coordinates | | | WM regions | Side | Voxels | Total voxels | Mean *p* |
|---|---|---|---|---|---|---|---|
| X | Y | Z | | | | | |
| -4 | 19 | 17 | | | | 45,922 | $p_{min}$ = .009 |
| | | | Corpus callosum | B | 2,066 | 3,696 | .018 |
| | | | | G | 1,257 | | |
| | | | | S | 373 | | |
| | | | Anterior corona radiata | R | 1,182 | 2,166 | .020 |
| | | | | L | 984 | | |
| | | | Superior longitudinal fasciculus | R | 753 | 1,547 | .021 |
| | | | | L | 794 | | |
| | | | External capsule | R | 576 | 1,210 | .019 |
| | | | | L | 634 | | |
| | | | Superior corona radiata | R | 630 | 937 | .027 |
| | | | | L | 307 | | |
| | | | Posterior corona radiata | R | 370 | 634 | .024 |
| | | | | L | 264 | | |
| | | | Anterior limb of internal capsule | R | 375 | 624 | .020 |
| | | | | L | 249 | | |
| | | | Retrolenticular part of internal capsule | R | 246 | 480 | .023 |
| | | | | L | 234 | | |
| | | | Sagittal stratum | R | 236 | 414 | .024 |
| | | | | L | 178 | | |
| | | | Posterior thalamic radiation | R | 169 | 334 | .034 |
| | | | | L | 165 | | |
| | | | Fornix | R | 95 | 152 | .030 |
| | | | | L | 57 | | |
| | | | Uncinate fasciculus | R | 78 | 117 | .019 |
| | | | | L | 39 | | |

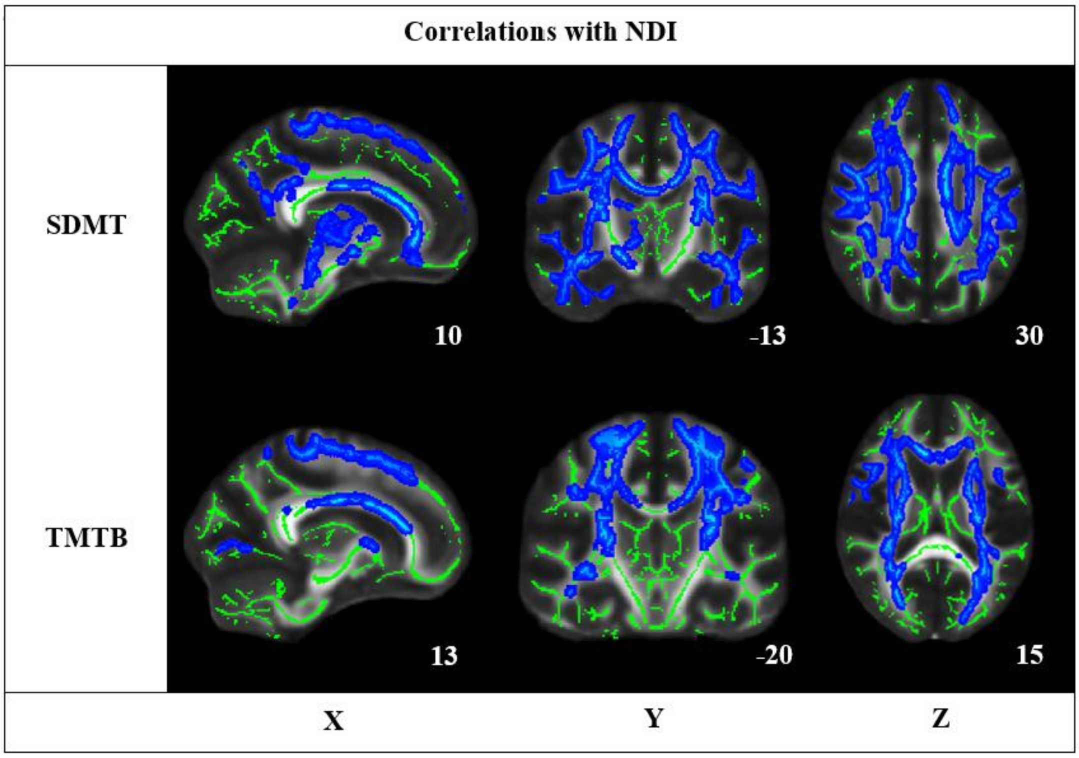


**Fig. S1** WM regions with significant clusters of decreased NDI with decreasing neuropsychological performance (blue; not controlled for UWDRS-N scores) in WD patients are shown on the study-specific mean WM skeleton (green) on the FMRIB58 FA template (TFCE, FWE-corrected $p \leq .05$). Results are presented on three representative slices (in MNI coordinates). The TBSS fill script was used to aid visualization. The images are displayed in accordance with radiological convention, meaning that the left side of the images corresponds to the right hemisphere of the brain and vice versa

**Table S10 WM regions where NDI decreased with poorer SDMT performance**

| Coordinates | | | WM regions | Side | Voxels | Total voxels | Mean *p* |
|---|---|---|---|---|---|---|---|
| X | Y | Z | | | | | |
| 42 | -5 | -29 | | | | 39,596 | $p_{min}$ = .025 |
| | | | Corpus callosum | B | 1,623 | 3,782 | .035 |
| | | | | G | 1,194 | | |
| | | | | S | 965 | | |
| | | | Superior corona radiata | R | 914 | 1,732 | .038 |
| | | | | L | 818 | | |
| | | | External capsule | R | 920 | 1,591 | .039 |
| | | | | L | 671 | | |
| | | | Anterior corona radiata | R | 981 | 1,451 | .037 |
| | | | | L | 470 | | |
| | | | Superior longitudinal fasciculus | R | 585 | 1,107 | .037 |
| | | | | L | 522 | | |

| | | | | | | | |
|---|---|---|---|---|---|---|---|
| | | | Posterior thalamic radiation | R | 266 | 830 | .037 |
| | | | | L | 564 | | |
| | | | Posterior corona radiata | R | 455 | 787 | .036 |
| | | | | L | 332 | | |
| | | | Anterior limb of internal capsule | R | 389 | 651 | .040 |
| | | | | L | 262 | | |
| | | | Sagittal stratum | R | 430 | 649 | .035 |
| | | | | L | 219 | | |
| | | | Retrolenticular part of internal capsule | R | 323 | 456 | .037 |
| | | | | L | 133 | | |
| | | | Posterior limb of internal capsule | R | 191 | 361 | .042 |
| | | | | L | 170 | | |
| | | | Superior cerebellar peduncle | R | 116 | 198 | .045 |
| | | | | L | 82 | | |
| | | | Fornix | R | 174 | 187 | .039 |
| | | | | L | 13 | | |
| | | | Cerebral peduncles | R | 157 | 175 | .048 |
| | | | | L | 18 | | |
| | | | Pontine crossing tract | | | 154 | .045 |
| | | | Medial lemniscus | R | 68 | 128 | .046 |
| | | | | L | 60 | | |

**Table S11 WM regions where NDI decreased with poorer TMTB performance**

| Coordinates | | | WM regions | Side | Voxels | Total voxels | Mean *p* |
|---|---|---|---|---|---|---|---|
| X | Y | Z | | | | | |
| -35 | -26 | 34 | | | | 27,456 | $p_{min}$ = .010 |
| | | | Corpus callosum | B | 1,769 | 2,540 | .022 |
| | | | | G | 461 | | |
| | | | | S | 310 | | |
| | | | Superior corona radiata | R | 1,052 | 2,115 | .019 |
| | | | | L | 1,063 | | |
| | | | Anterior corona radiata | R | 908 | 1,397 | .022 |
| | | | | L | 489 | | |
| | | | External capsule | R | 717 | 1,303 | .034 |
| | | | | L | 586 | | |
| | | | Superior longitudinal fasciculus | R | 508 | 1,114 | .024 |
| | | | | L | 606 | | |
| | | | Posterior thalamic radiation | R | 361 | 883 | .025 |
| | | | | L | 522 | | |
| | | | Posterior corona radiata | R | 493 | 758 | .021 |
| | | | | L | 265 | | |
| | | | | R | 343 | 640 | .023 |

| | | | | | | | |
|---|---|---|---|---|---|---|---|
| | | | Anterior limb of internal capsule | L | 297 | | |
| | | | Posterior limb of internal capsule | R | 298 | 534 | .024 |
| | | | | L | 236 | | |
| | | | Retrolenticular part of internal capsule | R | 282 | 410 | .029 |
| | | | | L | 128 | | |
| | | | Superior fronto-occipital fasciculus | R | 94 | 154 | .019 |
| | | | | L | 60 | | |
| | | | Sagittal stratum | R | 86 | 136 | .041 |
| | | | | L | 50 | | |

#### 2.2 Correlations between FA and clinical scores

##### Table S12 WM regions where FA decreased with higher UWDRS-N scores

| **Coordinates** | | | **WM regions** | **Side** | **Voxels** | **Total voxels** | **Mean *p*** |
|---|---|---|---|---|---|---|---|
| **X** | **Y** | **Z** | | | | | |
| -17 | 22 | 25 | | | | 13,341 | $p_{min}$ = .028 |
| | | | Corpus callosum | B | 1,304 | 1,997 | .034 |
| | | | | G | 641 | | |
| | | | | S | 52 | | |
| | | | Anterior corona radiata | R | 708 | 1,439 | .035 |
| | | | | L | 731 | | |
| | | | External capsule | L | | 746 | .043 |
| | | | Superior corona radiata | R | 370 | 559 | .034 |
| | | | | L | 189 | | |
| | | | Superior longitudinal fasciculus | L | 525 | 525 | .034 |
| | | | Medial lemniscus | R | 114 | 248 | .040 |
| | | | | L | 134 | | |
| | | | Superior cerebellar peduncle | R | 146 | 237 | .040 |
| | | | | L | 91 | | |
| -26 | -74 | 1 | | | | 204 | $p_{min}$ = .046 |
| -30 | -48 | 16 | | | | 125 | $p_{min}$ = .047 |
| | | | Posterior thalamic radiation | L | 103 | 103 | .048 |

**Table S13 WM regions where FA decreased with poorer TMTA performance (controlled for UWDRS-N scores)**

| Coordinates | | | WM regions | Side | Voxels | Total voxels | Mean *p* |
|---|---|---|---|---|---|---|---|
| X | Y | Z | | | | | |
| 5 | -34 | -26 | | | | 16,781 | $p_{min}$ = .019 |
| | | | Corpus callosum | B | 1,383 | 2,341 | .035 |
| | | | | G | 557 | | |
| | | | | S | 401 | | |
| | | | Anterior corona radiata | R | 814 | 1,804 | .034 |
| | | | | L | 990 | | |
| | | | External capsule | R | 430 | 1,114 | .035 |
| | | | | L | 684 | | |
| | | | Superior corona radiata | R | 260 | 659 | .035 |
| | | | | L | 399 | | |
| | | | Posterior thalamic radiation | L | 599 | 599 | .042 |
| | | | Cerebral peduncles | R | 105 | 449 | .029 |
| | | | | L | 344 | | |
| | | | Superior cerebellar peduncles | R | 185 | 362 | .028 |
| | | | | L | 177 | | |
| | | | Fornix | R | 161 | 349 | .035 |
| | | | | L | 188 | | |
| | | | Retrolenticular part of internal capsule | R | 93 | 305 | .035 |
| | | | | L | 212 | | |
| | | | Posterior limb of internal capsule | R | 86 | 269 | .032 |
| | | | | L | 183 | | |
| | | | Medial lemniscus | R | 98 | 236 | .028 |
| | | | | L | 138 | | |
| | | | Cingulum | L | 214 | 214 | .035 |
| | | | Sagittal stratum | R | 21 | 208 | .035 |
| | | | | L | 187 | | |
| | | | Posterior corona radiata | R | 107 | 188 | .037 |
| | | | | L | 81 | | |
| | | | Corticospinal tract | L | 135 | 135 | .028 |
| | | | Pontine crossing tract | | | 125 | .030 |
| | | | Anterior limb of internal capsule | R | 32 | 117 | .039 |
| | | | | L | 85 | | |

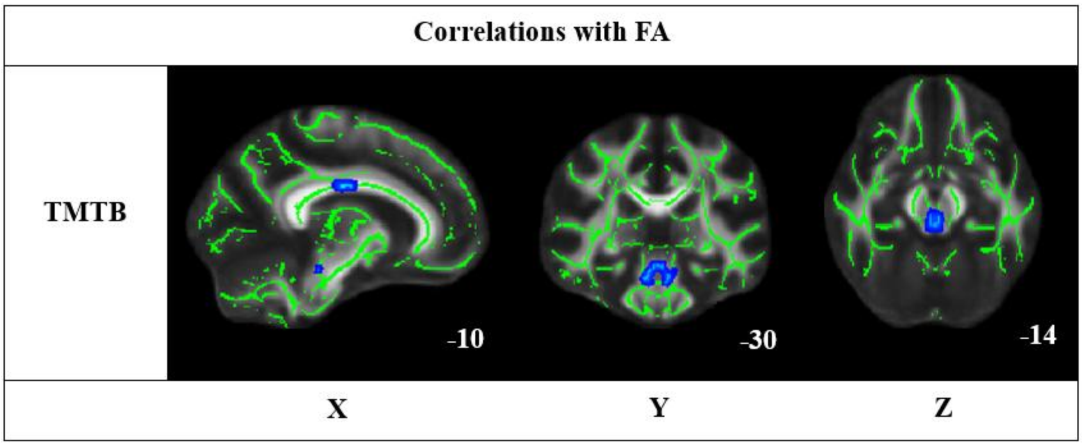


**Fig. S2** WM regions with significant clusters of decreased FA with decreasing TMTB performance (blue; not controlled for UWDRS-N scores) in WD patients are shown on the study-specific mean WM skeleton (green) on the FMRIB58 FA template (TFCE, FWE-corrected $p \leq .05$). Results are presented on three representative slices (in MNI coordinates). The TBSS fill script was used to aid visualization. The images are displayed in accordance with radiological convention, meaning that the left side of the images corresponds to the right hemisphere of the brain and vice versa

**Table S14 WM regions where FA decreased with poorer TMTB performance (not controlled for UWDRS-N scores)**

| Coordinates | | | WM regions | Side | Voxels | Total voxels | Mean *p* |
|---|---|---|---|---|---|---|---|
| X | Y | Z | | | | | |
| 1 | -30 | -14 | | | | 288 | $p_{min} = .039$ |
| -11 | -16 | 29 | | | | 208 | $p_{min} = .042$ |
| | | | Corpus callosum | B | 204 | 204 | .047 |
| 12 | -17 | 30 | | | | 176 | $p_{min} = .040$ |
| | | | Corpus callosum | B | 163 | 163 | .045 |
